# RNAsum: a tool for personalised genome and transcriptome interpretation for improved cancer diagnostics

**DOI:** 10.1101/2025.01.10.24319650

**Authors:** Sehrish Kanwal, Jacek Marzec, Joseph H.A. Vissers, Peter Diakumis, Leila N. Varghese, Luke Tork, Kym Pham Stewart, Oliver Hofmann, Stephen J. Luen, Sean M. Grimmond

## Abstract

**Background:** The integration of whole-genome sequencing (WGS) and whole-transcriptome sequencing (WTS) has revolutionized cancer diagnostics by enabling comprehensive molecular profiling of tumours. WGS uncovers genomic alterations such as single nucleotide variants, structural variants, and copy number changes, whereas WTS reveals their functional consequences through expression profiles and fusion detection. Together, these technologies offer unparalleled potential to guide precision oncology by identifying actionable biomarkers and stratifying patients for targeted therapies or clinical trials. Recent studies have shown that nearly half of patients experience improved clinical outcomes when treatment is guided by combined WGTS analysis. However, dedicated tools for automated single-patient integration and reporting of WGTS data within routine clinical bioinformatics workflows remain limited, representing a significant gap in personalised cancer sample interpretation.

**Results:** We developed RNAsum, an open-source tool for integrating and interpreting whole-genome sequencing (WGS) and whole-transcriptome sequencing (WTS) data from individual cancer patient samples. RNAsum compares patient data to The Cancer Genome Atlas (TCGA) cohorts, integrating quantitative expression data with genomic findings to corroborate and prioritise clinically relevant alterations and enhance diagnostic accuracy. Clinical applicability evaluation performed across 60 patients demonstrated that 68% (140/205) of the clinically reportable variants identified by WGS, spanning copy number changes, truncating mutations and gene fusions, were supported at the RNA level as detected by WTS and reported by RNAsum. Case studies further highlight the ability of RNAsum to support clinically reportable variants, refine diagnoses, and identify novel therapeutic targets, particularly in complex cases involving multiple genomic alterations and drug resistance mechanisms.

**Conclusion:** RNAsum effectively bridges the gap between genome and transcriptome analyses, significantly advancing the integration of multiomics data in personalized cancer care. Its ability to corroborate clinically reportable variants at the RNA level and elucidate complex alterations, including those driving drug resistance, highlights RNAsum’s potential to improve molecular tumour profiling and support clinical decision-making. Freely available as an R package on GitHub at https://github.com/umccr/RNAsum, RNAsum provides an accessible and scalable solution for researchers and clinicians, representing a significant step towards the routine application of integrated genomic and transcriptomic analyses in precision oncology.

## Background

High-throughput sequencing (HTS) technologies have revolutionized the field of precision oncology by enabling fast and accurate analysis of whole-genome and transcriptome data from tumours [1,2]. Owing to the decreasing sequencing cost, whole-genome sequencing (WGS) provides valuable insights into the molecular mechanisms underlying oncogenesis and genomic aberrations such as single nucleotide variants (SNVs), small insertions and deletions (indels), structural variants (SVs) and copy-number variants (CNVs) [3,4]. Furthermore, it has shown great utility in studying clinically actionable cancer genome alterations including tumour mutational burden (TMB) [5], homologous recombination deficiency (HRD) [6] and microsatellite instability (MSI) [7].

As the field of HTS continues to expand and support precision oncology initiatives globally, whole-transcriptome sequencing (WTS) has gained immense traction in cancer research and personalized medicine [8] due to its ability to provide comprehensive insights into the transcriptome, the complete set of gene readouts called RNA molecules in a cell or tissue. It allows quantification of expression levels to understand gene activation or silencing in a cancerous cell [9].

Combined whole-genome and transcriptome sequencing (WGTS) has demonstrated the ability to provide a nearly complete fingerprint of patients’ tumour profiles and is increasingly being used to molecularly diagnose challenging malignancies [10–14]. While WGS provides insights into both coding and non-coding genomic alterations by detecting a range of variant types, including SNVs, indels, CNVs and SVs, integrating RNA sequencing (RNA-seq) with WGS enables confirmation of the functional consequences of these somatic changes. This comprehensive diagnostic approach can improve precision oncology practices by providing complete characterisation and molecular profiling for the identification of targetable alterations that stratify patients for eligible clinical trials and off-label drug treatment [15]. By understanding the unique genetic mutations and expression profile of a patient, clinicians can use these data for disease detection, prognosis and tailor therapies to be more effective and minimize side effects.

Recent studies have demonstrated the significant value of integrating RNA expression data with genomic profiling in precision oncology. [16] reported that nearly half of patients demonstrated positive clinical outcomes when treatment was guided by combined WGTS analysis. These findings highlighted how RNA data can confirm the functional consequences of genomic mutations, either independently or in combination with DNA sequencing, thereby informing treatment decisions. This approach lays the groundwork for future precision oncology efforts, including our current research, which aims to incorporate WGTS data into routine clinical care for cancer patients.

While current clinical genomics tests focus primarily on WGS data, integrated transcriptome analysis has shown significant value in precision oncology. Studies have demonstrated the importance of cross-validating WGS findings with transcriptome-based results [17,18]. However, translating these combined datasets into clinically actionable, patient-level findings within routine bioinformatics workflows remains non-trivial. To address this gap, we developed RNAsum, an automated report-centric tool that streamlines the integration of user-provided WGS and WTS data from individual patient samples with matched tumour-type reference cohorts. RNAsum enables comprehensive analysis of the transcriptional consequences of somatic events, compares patient data against The Cancer Genome Atlas (TCGA) [19] reference cohort, and generates self-contained interactive reports summarising key findings for use in precision oncology workflows and molecular tumour board discussions. We demonstrate the implementation and clinical utility of RNAsum through case studies, discuss its features and limitations, and provide detailed usage information, thereby establishing its value in routine clinical practice for WGTS interpretation in precision oncology.

### Implementation

RNAsum is an open-source R package that enables the integration and interpretation of WGS and WTS data from individual cancer patient samples while remaining easy to install and requiring minimal technical expertise or management overhead.

### Reference data

To assist in the interpretation of gene expression data prospectively generated from patient cancer samples, we applied a solution that allows the comparison of single-subject tumour-only expression profiles to large cancer-matched expression landscapes taken from the TCGA. Read count data from 10,079 cancer patients, covering 33 cancer types (Table S1) were obtained from the National Cancer Institute (NCI) Genomic Data Commons (GDC) Data Portal (https://portal.gdc.cancer.gov) via the GDC Application Programming Interface (API, https://gdc.cancer.gov/developers/gdc-application-programming-interface-api) and data release 40.0 (March 29, 2024, https://docs.gdc.cancer.gov/Data/Release_Notes/Data_Release_Notes/#data-release-400). These reference profiles were then harmonized with the individual sample to determine the relative gene expression level of any gene of interest in the individual, expressed as a percentile of the relative to the reference patient cohort expression spectrum, hereinafter referred to as “relative percentiles” (see the “WTS data integration” section for a detailed description).

Additionally, users can provide a customized list of genes of interest (GOIs) to the workflow, enabling exploration of their expression profiles in investigated patients relative to corresponding reference cohorts. If no custom gene set is supplied, the workflow defaults to analyse three preassembled gene sets: (1) “immune-related genes” (listed in the Immune_markers PanelApp gene panel, https://panelapp-aus.org/panels/243) and included to monitor potential cancer immune regulation), (2) “homologous recombination deficiency (HRD) genes” (listed in the Homologous_recombination_deficiency PanelApp gene panel, https://panelapp-aus.org/panels/242), and (3) “known cancer genes”, a curated set of 1,315 genes associated with tumorigenesis (defined at https://github.com/umccr/gene_panels/releases). These exemplar gene sets are routinely reviewed in individual patients to explore the expression of immune-related genes, genes associated with HRD, and key cancer-related genes.

Finally, we collected information from public knowledge bases, including FusionGDB [20], Cancer Genome Interpreter (CGI) [21], Clinical Interpretation of Variants in Cancer (CIViC) [22], Variant Interpretation for Cancer Consortium (VICC, https://cancervariants.org) and OncoKB [23,24], to annotate WGTS findings and to determine the clinical significance of the identified alterations.

### Workflow overview

The RNAsum workflow includes four data components (data from patient samples are summarized in Table 1) and consists of two integration and one annotation steps (Fig. 1). First, mRNA profiles from the patient and external (TCGA) reference cohort are combined at the WTS level (components 1 and 2). In the second step, the expression data and detected gene fusions are overlaid with genomic alterations at the WGTS level (component 3). Finally, clinically relevant annotations from public knowledge bases are added to facilitate interpretation (component 4).

**Fig. 1.**
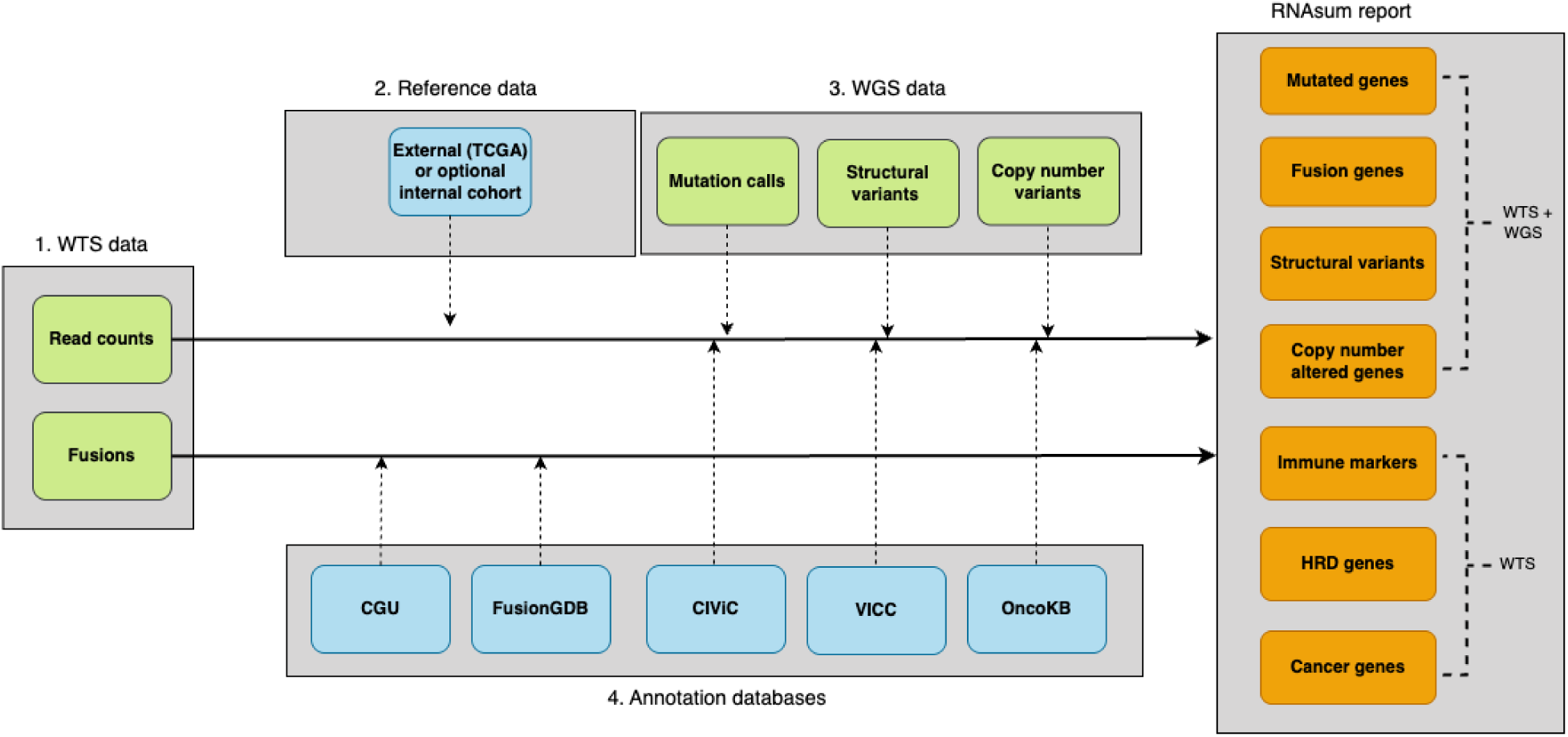
Overview of the integrated genome and transcriptome analysis workflow. The process is divided into four main stages: (1) read count and gene fusion data collection, (2) reference data processing, (3) integration with WGS-based data, and (4) results annotation. Colour coding is used to distinguish different elements: green represents inputs from a patient, blue indicates reference data, and orange denotes outputs. This workflow enables comprehensive analysis of genomic and transcriptomic data by combining multiple data types and reference sources.

**Table 1.** Summary of input data from a patient (components in green in Fig. 1).

| Workflow component (platform) | Input data | Data source | Required | Example tool/database |
| --- | --- | --- | --- | --- |
| **1** (WTS) | Transcript/gene abundances | Patient | Yes | Salmon [25], Kallisto [26] |
| **1** (WTS) | Fusion genes | Patient | No | Arriba [27], DRAGEN [28] |
| **3** (WGS) | SNVs/indels | Patient | No | PCGR [29] |
| **3** (WGS) | CNVs | Patient | No | PURPLE [30] |
| **3** (WGS) | SVs | Patient | No | Manta [31] |

#### WTS data integration

The read count data from two sources are used as an inputs for the integration at the WTS level: (1) the patient sample and (2) an external reference cohort, which may either match the patient’s cancer type (tumour-specific) or consist of all patients available within TCGA (pan-cancer) (see the “Reference data collection” section and Table S1). Tumour-specific cohorts offer more context-relevant comparisons, whereas the pan-cancer cohort provides larger sample sizes and broader expression ranges, improving percentile resolution, especially for genes expressed across multiple tumour types. The pan-cancer cohort is especially useful in cases where the tumour origin is unknown, the histology is rare or ambiguous, or where broader benchmarking across cancer types is of interest.

Users can also choose between two TCGA reference cohort modes: the “full” or a memory-efficient “representative” subset. While the “full” mode improves precision, the default “representative” mode supports better scalability and integration into routine workflows, particularly when a memory-demanding pan-cancer reference cohort is used. To evaluate the impact of the two modes, we analysed 10 of the 60 patient cases used in the RNAsum clinical utility evaluation (see “Clinical applicability evaluation”). Specifically, we examined five cases combined with the tumour-matched cohort and five cases combined with the pan-cancer TCGA reference cohort. The benchmarking results revealed that 99% of the “known cancer genes” (see “Reference data collection”) presented <10 percentile point differences between the two modes (Fig. S1), with discrepancies affecting mainly genes with low or undetectable expression. This reflects typical behaviour in transcriptomics data, where low-abundance transcripts are more susceptible to variation after normalization.

Users should note that the choice of reference cohort can substantially affect the interpretation of expression percentile rankings. Pan-cancer cohort provides broad context but may dilute signals for genes with tissue-specific expression patterns. For histology-specific driver genes, for example, genes with tissue-restricted overexpression such as AR in prostate cancer or CDX2 in colorectal cancer, a matched tumour-type cohort (e.g., TCGA-PRAD or TCGA-COAD) may be more appropriate. RNAsum supports user-specified cohort selection via the --dataset parameter. The default pan-cancer cohort is recommended as a first-pass analysis, with tumour-type-specific cohorts as a confirmatory step for histology-specific genes.

We evaluated the impact of batch effect correction using an internal reference cohort (Fig. S2). While comparisons between clinical RNA-seq data and the TCGA reference cohort are expected to exhibit prominent batch effects, arising from differences in library preparation chemistry (ribo-depletion versus poly(A) selection), sequencing platforms, and centre-specific processing, our evaluation used a clinically focused assessment approach distinct from standard PCA-based methods. Unlike standard batch assessments that use top-variable genes, which, as the most variable features in the dataset, are expected to reflect both biological heterogeneity and any systematic technical differences in library preparation chemistry, our evaluation restricted the gene set to those with WGS-detected clinical alterations: truncating SNVs, rearrangements, copy number losses, and amplifications. Batch effect correction was applied to all patients in this cohort. The assessment was performed across 10 representative cases (five combined with tumour-matched and five with pan-cancer reference cohorts) and included all clinically reported variants predicted to impact RNA expression. Using this clinically relevant evaluation framework, batch effect correction had limited impact on the variant prioritisation outcomes reported here. RNAsum provides the --batch_rm parameter and dedicated batch assessment functions (assess_batch_effects(), quick_batch_check()) to enable users to evaluate and correct batch effects in their own datasets. We recommend their use particularly when clinical library preparation protocols differ substantially from the TCGA cohort.

The input read count data are combined and transformed to counts per million (CPM) values, genes with CPM > 1 in fewer than 10% of samples are excluded, and then the data are normalized to account for sample-specific effects (Fig. S3). Prior to Z-score computation, expression values are log2-transformed [log2(CPM + 0.25)], which stabilises variance, reduces the influence of extreme values, and produces an approximately normal distribution suitable for parametric statistics, an approach standard in tools such as limma-voom [32] and edgeR [33]. Gene expression levels in the patient sample are reported as percentiles relative to the expression spectrum of the reference patient cohort. Additionally, WTS data are standardized using Z-scores, which express deviations from the mean in terms of standard deviations. This transformation enables direct comparison across samples and facilitates the interpretation of expression values (Fig. S4).

To characterise the strandedness of the TCGA reference data used by RNAsum, we performed a pan-cancer census of all 11,505 TCGA samples available through the NCI Genomic Data Commons (GDC, Data Release 40.0), confirming that 32 of 33 TCGA cancer projects (96.6%; n = 11,114 samples) use non-strand-specific library preparation, consistent with GDC harmonisation policy. RNAsum accordingly uses unstranded GDC counts for all reference cohort expression metrics (detailed methods in Supplementary Note 1). When patient WTS data are generated using strand-specific protocols, a structural analysis of GENCODE v36 identified 21 clinically actionable cancer genes with ≥10,000 bp antisense overlap, including *ALK*, *BRCA1*, *CDKN2A*, *NF1*, *RB1*, *EGFR*, and *ERBB2*, that may exhibit non-trivial percentile displacement relative to the unstranded reference. These genes are flagged in the RNAsum HTML report with a recommendation for additional interpretive caution.

RNA-only mode is intended for situations where WGS data are unavailable or have not yet been processed - for example, when rapid turnaround of transcriptomic findings is required ahead of genome sequencing results, when WGS was not clinically indicated, or when RNAsum is being applied retrospectively to archived WTS data from cohorts without matched WGS. In this mode, RNAsum still provides expression profiling, outlier detection, fusion gene identification, and comparison against TCGA reference cohorts, but without the variant prioritisation and WGS-WTS integration features that require processed WGS data as input. This mode may also be useful for research applications where WGS data are unavailable, such as re-analysis of publicly available WTS datasets.

#### WGTS data integration

WTS data for each patient are integrated with matched WGS data by overlaying gene expression levels onto genes harbouring genomic alterations. Various tools can provide the necessary input files in compatible formats, including SNVs and indels from tools such as the Personal Cancer Genome Reporter (PCGR) [29], SVs from callers like MANTA [31], and CNVs from tools such as PURPLE purity ploidy estimator [30] (Fig. 1). Moreover, gene fusions identified by Arriba [27] or DRAGEN [28] and supporting WGS-derived SVs are prioritised, as this concordant evidence strengthens their functional relevance. To enhance usability, RNAsum accepts simplified TSV input files containing gene-level SV or CN information. This flexibility enables users to integrate results generated by their preferred analysis tools. The content presented in the output tables is drawn directly from the user-provided TSVs, ensuring compatibility with diverse analytical workflows and eliminating dependence on any specific software. Gene-level integration between WTS and WGS data layers uses Ensembl IDs for expression matrix merging and HGNC symbols for variant overlay. Consistent annotation versions across upstream tools are therefore recommended to minimise the risk of unmatched genes in the variant expression overlay.

Genome-based findings, if provided, serve as the primary source for prioritising expression profiles, ensuring that the differences in expression levels between patient sample and the corresponding reference cohort for key genes are evaluated first. The integrated WTS and WGS data are then ranked by the observed difference, highlighting genes with the greatest potential clinical impact. All reported alterations are annotated with interpretation-relevant information from public knowledge bases, including CGI [21], VICC (https://cancervariants.org), OncoKB [23,24] and CIViC [22]. Additionally, detected gene fusions are functionally annotated with information from FusionGDB [20].

### WGTS variant interpretation

The outputs of the pipeline are an interactive report that prioritises clinically relevant information to facilitate results interpretation, as well as supplementary resources that can be used programmatically for further data exploration and comparisons against other patient cohorts. This report includes searchable tables and plots organised into several sections detailed below and illustrated in Fig. 2 (see the Case study #1 section for more details). Each section features an interactive “summary” table and “expression profiles” plots.

**Fig. 2.**
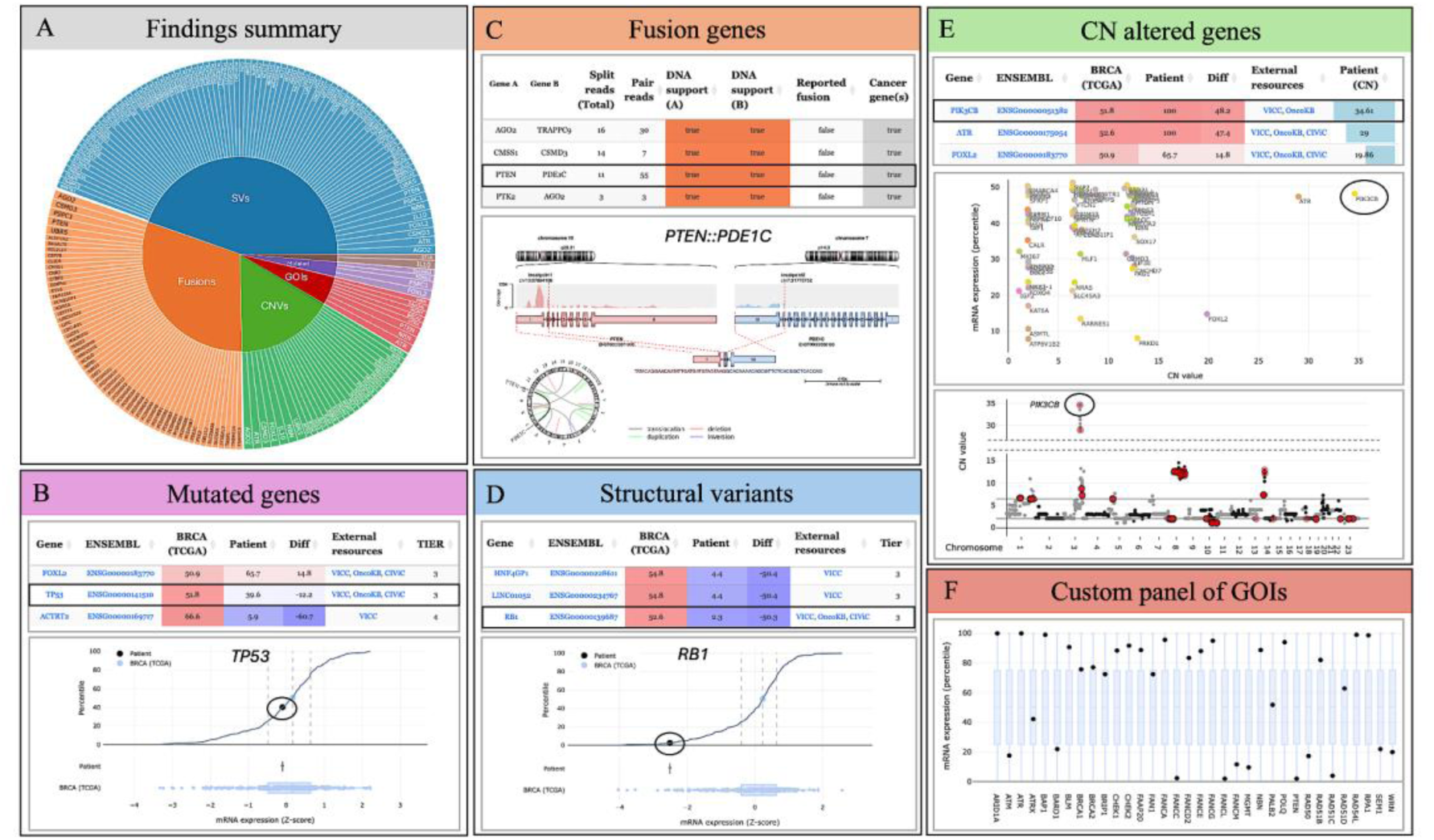
Overview of RNAsum interactive report structure and visualisation features. The diagram illustrates the organisation of the report into key sections, including the input data summary, findings summary (**A**), mutated genes (**B**), fusion genes (**C**), structural variants (**D**), CN altered genes (**E**), and a custom panel of genes of interest (**F**), each described in detail in the main text. Example plots and tables demonstrate how RNAsum prioritises and visualises clinically relevant findings to support interpretation The expression values presented in the “summary” tables are presented as percentiles, and “expression profiles” plots are shown as Z-scores. *GOIs — genes of interest*.

The “summary” table contains comprehensive information about individual genes’ mRNA expression levels in a patient sample, the average mRNA expression across samples from TCGA cancer patients, and the difference between these two measurements. Notably, mRNA measurements are presented in both percentiles, to facilitate intuitive interpretation, and Z-scores. Genes in these tables are sorted based on section-specific criteria and then by the decreasing absolute values representing the difference between patient and TCGA expression levels. The “summary” table also includes hyperlinks to relevant knowledge bases, such as VICC (https://cancervariants.org), OncoKB [23,24] and CIViC [22], providing additional context, as well as information specific to each report section. The interactive plots comprise two main components: a distribution panel showing percentile values as a function of expression levels and a box plot panel displaying expression levels for each gene in the patient sample compared with the reference cancer cohort. Additionally, we included a bar plot illustrating the read counts of individual genes across all samples.

### Input data summary

The WTS input data are summarized at the top of the report, providing a comprehensive overview of the analysis. This section includes information about reference cohorts and patient samples used in the analysis, details about included and excluded genes along with the reasoning behind these decisions, visual representations of the initial library size generated across all samples, read count data filtering, transformation and normalization. Moreover, it contains interactive exploratory data analysis plots, including principal component analysis (PCA) and relative log expression (RLE) plots, which facilitate the identification of key factors affecting variability in the expression data.

### Findings summary

The findings from the WGTS data integration are summarized in the form of an interactive doughnut plot and table presenting genes with detected alterations, which are further detailed in the corresponding report sections (Fig. 2A). The genes are ordered by the number of report sections in which they appear, highlighting those of particular interest owing to evidence from multiple sources. Moreover, all affected genes are hyperlinked to databases, including VICC (https://cancervariants.org), OncoKB [23,24] and CIViC [22], providing additional evidence for their clinical significance.

### Mutated genes

The integrated WGTS data for genes with detected SNVs or indels are presented in an interactive “summary” table and “expression profiles” plots, as described previously (Fig. 2B). The “summary” table provides detailed information on mRNA levels for individual genes, along with detected variant tiers, variant characteristics (such as type, class, and allele frequency), and associated proteomic changes. Genes in the table are primarily ordered by variant tier, reflecting their classification and clinical relevance, and secondarily by the percentile difference between the patient’s gene expression levels and those observed in the TCGA dataset.

### Fusion genes

Gene fusions detected in WTS are summarized in an interactive table that provides comprehensive information, including the number of supporting split reads, the genomic coordinates of the fused genes, the locations of the breakpoints, and whether they overlap with structural events identified in WGS data (Fig. 2C). Gene fusions that support WGS-based SVs or are reported in FusionGDB [20] or CGI [21] databases are prioritised, given the additional evidence for their functional relevance and potential clinical significance in the investigated patient. These fusions are presented in a genomic context via a Circos plot, followed by an interactive “summary” table and “expression profiles” plots that demonstrate the mRNA levels of individual fusion genes observed in both the patient and reference cohorts. Additionally, this section provides visualisations (generated by Arriba [27]) that offer detailed information about the transcripts involved in the predicted fusions. These visualisations include the orientation of the transcripts, the retained exons in the fusion transcript, and statistics about the number of supporting reads.

### Structural variants

The integrated WGTS data for genes affected by other SVs detected in the WGS are presented in the “summary” table ordered by the SV score and the difference between the investigated patient and TCGA expression levels (Fig. 2D). In addition to the genomic information associated with detected SVs, such as genomic location and CN changes, the table provides details about affected transcripts and potential overlapping gene fusions. The accompanying “expression profiles” plots display the expression levels of the affected genes as measured in the patient samples and reference cohort samples.

### Copy number altered genes

The “CN altered genes” section overlays mRNA expression data with per-gene somatic CN data and information about SNVs and indels (Fig. 2E). It begins with an interactive Manhattan-like plot that presents CN values for genes located within gained or lost regions. Additionally, CNs and expression levels of affected genes are displayed in a scatterplot, which shows the per-gene difference in mRNA expression between the patient sample and the reference cohort as a function of the corresponding CN values. Following these visualisations, the section includes a “summary” table, ordered by the CN value and the difference between the investigated patient and TCGA expression levels, complemented by information derived from the “Mutated genes” section. The “expression profiles” plots depict the expression levels of the affected genes, as measured in the patient and reference cohort samples. These combined visualisations and data tables allow for the correlation of gene expression with CN alterations, providing insights into their potential clinical significance.

### Custom panel of genes of interest

The final section of the report presents results derived exclusively from WTS data and focuses on the expression profiles of a customizable list of GOIs provided by the user or three specific sets of genes: (1) “immune-related genes”, (2) “HRD genes” and (3) “known cancer genes” (Fig. 2F). The mRNA levels of genes involved in these gene sets are presented in the corresponding “summary” tables and “expression overview” box plots.

## Results

### Clinical applicability evaluation

To assess the clinical utility of RNAsum in interpreting aberrant events detected in WGS, we reviewed 81 patient cases based on the following criteria: 1) availability of matched WGS and WTS data, 2) ≥1 somatic genetic alteration was detected by WGS and clinically reported and 3) RNAsum corroborated the predicted impact on RNA expression of ≥1 clinically reported genetic alteration. Two cases failed criterion 2, due to insufficient tumour cellularity. In 19 cases, no genetic alterations were reported that impacted RNA expression, failing criterion 3. Of the remaining 60 cases, all clinically reported variants predicted to impact RNA expression were aggregated. These included truncating SNVs and small indels, inactivating rearrangements, CN losses (low expression), amplifications (high expression) and fusions (fusion mRNA expression).

Across all clinically reportable variants identified by WGS, 205 were classified as variant types potentially amenable to qualitative or quantitative support from WTS data. Among these, 140 (68%) variants were supported at the RNA level at a top/bottom quartile threshold (≥75th or ≤25th percentile relative to the TCGA reference cohort), as detected by WTS and reported by RNAsum (Fig. 3). To assess the robustness of this finding to threshold choice, we reanalysed WTS support using 10-percentile bins across progressively stricter thresholds. At the ≥90th/≤10th percentile threshold, 78 of 163 expression-assessed variants (47.6%) retained WTS support, compared with 10% expected under a null uniform distribution (exact one-sided binomial test, p = 1.6×10⁻³⁴), demonstrating that the observed corroboration rate is robust to threshold stringency and is not inflated by the choice of a permissive quartile cutoff (Fig. S5). The per-category analysis revealed an informative pattern across inactivating rearrangement subgroups. Inactivating rearrangements with a second hit showed markedly low expression, consistent with the expected loss of transcriptional output when both alleles are compromised. In contrast, inactivating rearrangements without a second hit did not show uniformly low expression, which is also expected - the presence of an intact wild-type allele maintains overall transcript abundance despite functional disruption of the rearranged copy. This finding underscores that expression level alone is not a sufficient readout for inactivating rearrangements without a second hit, and that the presence or absence of a second hit is an important biological consideration when interpreting WTS-based expression support for this variant class. According to the evaluated reports, WTS supported more than half of the clinically reportable variants in 52 out of 60 cases and all reportable variants in 31 cases, highlighting its complementary value to WGS (Table S2 and Fig. S6).

**Fig. 3.**
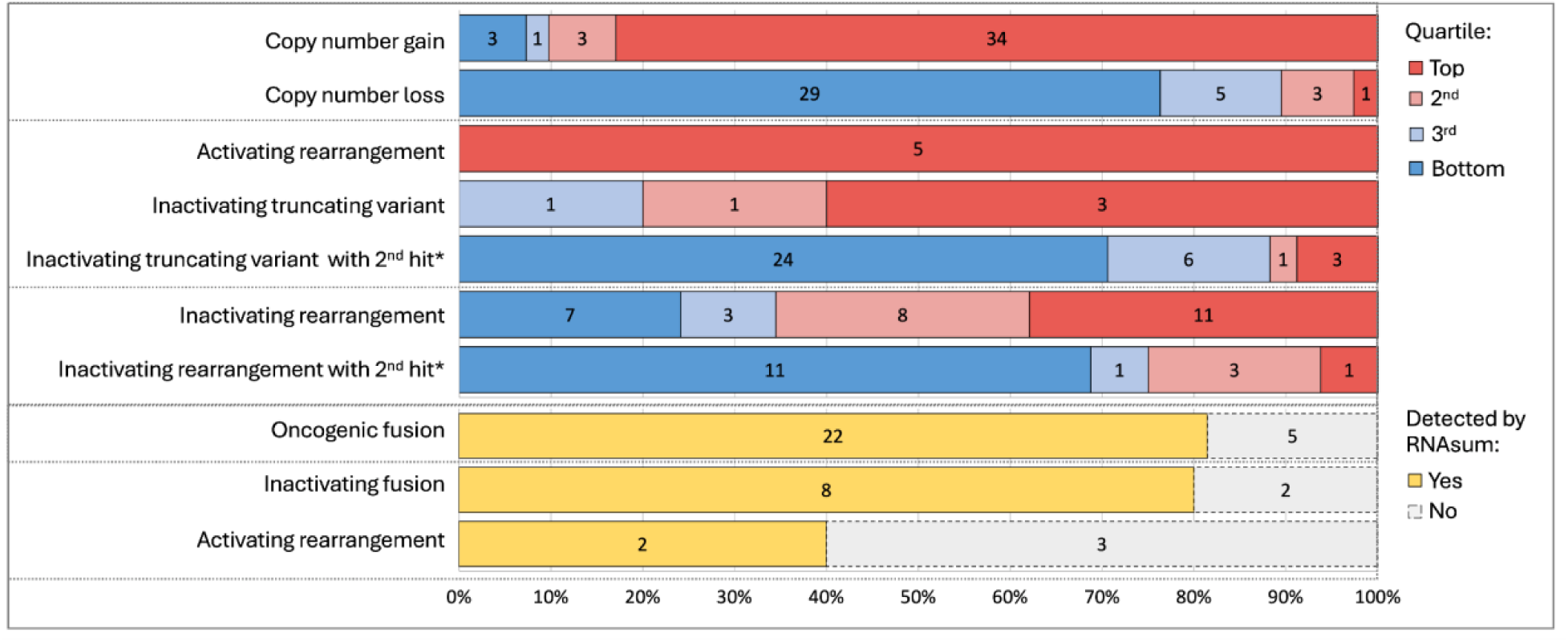
Transcriptomic corroboration of clinically relevant genomic alterations through RNAsum. The bar plot summarises the genomic alterations reported across 60 cases, categorised by WTS-based evidence, displayed using expression quartiles relative to the matched TCGA reference cohort (top quartile: ≥75th percentile; bottom quartile: ≤25th percentile). The stacked segments within each bar indicate the type of WTS support: relative expression quartiles for copy number alterations, activating and inactivating variants (with or without a second hit*), and detection of aberrant transcripts for structural rearrangements. The numerical labels within the bars indicate the number of variants supported by each WTS evidence category. This overview highlights the value of integrating WGS and WTS data to corroborate clinically reportable genomic variants, demonstrating RNAsum’s role in contextualising expression changes and fusion events alongside genomic alterations. A finer-resolution view of the same data using 10-percentile bins is provided in Fig. S5. *\*Includes inactivating variants on the X chromosome in males*.

These events included truncating mutations (nonsense and frameshifts), CN gains and losses, structural rearrangements, and gene fusions. WTS revealed high relative expression in 83% of CN gain variants (Fig. S7) at the quartile threshold (Fig. 3) and 68% at the ≥90th percentile threshold (Fig. S5); for CN loss variants, low relative expression (Fig. S7) was observed in 76% at the quartile threshold and 58% at the ≤10th percentile threshold. Both rates remain highly significant compared with the 10% expected under a null distribution (CN gains at ≥90th percentile: 28/41, p = 4.7×10⁻¹⁹; CN losses at ≤10th percentile: 22/38, p = 4.5×10⁻¹³; binomial tests). Additionally, bottom quartile relative expression was observed in 71% of inactivating truncating variants with a second hit and 69% of inactivating rearrangements with a second hit, where a second hit was defined as inactivating mutation of the second allele or loss of heterozygosity. In contrast, inactivating rearrangements without a second hit showed a markedly different expression distribution, with a median expression at the 73rd percentile, skewing high rather than low. This is consistent with a biological decoupling of transcription and function in this variant class: structural rearrangements that disrupt protein function do not necessarily silence transcription, and residual expression from the intact wild-type allele further elevates gene-level counts. A similar, though less pronounced, pattern is observed in inactivating truncating variants without a second hit. In addition, 76% of fusions and activating gene rearrangements were detected by RNAsum, including 81% of oncogenic fusions.

Although formal clinical follow-up was beyond the scope of this study, we identified 8 cases (13%) in which WTS evidence was essential to inform variant interpretation (Table S3). These included cases where promoter swap events were identified through outlier gene expression, where complex structural rearrangements predicted by WGS were confirmed as in-frame fusion transcripts by WTS, and where fusion gene detection informed diagnosis or predicted treatment resistance. These cases highlight a distinct role for WTS beyond corroboration, resolving genomic complexity and providing transcriptional evidence that directly influenced clinical interpretation. This comprehensive review of clinical cases underscores RNAsum’s effectiveness in detecting and contextualising clinically relevant genomic alterations, reinforcing its utility as a robust tool for curating clinical data.

### Case studies

To further demonstrate the utility of RNAsum in the clinical interpretation of somatic genomic alterations, we present two cases in which the data curated and summarized by RNAsum played a crucial role in guiding clinically significant decisions.

#### Case study #1

Whole-genome and whole-transcriptome sequencing was performed on a fresh metastatic triple-negative breast cancer (TNBC) biopsy sample (Table S2, Case 44). The tumour had a low tumour mutational burden (1.9 mutations/Mb) and contained somatic features typical of this disease [34]: a *TP53* truncating mutation (*TP53* c.637C>T p.R213*) accompanied by CN neutral loss of heterozygosity (LOH), chromosomal rearrangements of *PTEN* and *RB1,* and amplification of several genomic regions including chr8q.

RNAsum was run for this case using the TCGA breast cancer reference cohort. The expression of the *TP53* c.637C>T nonsense mutant allele was found to be transcriptionally active; manual inspection of RNA-seq alignments in IGV revealed 47% of 217 reads spanning the mutation site carried the variant, and RNAsum estimated the overall *TP53* expression level at the 41st percentile (Fig. 4A). These observations support the expression of a truncated p53 open reading frame and its interpretation as an LOF mutation [35,36]. The WGS-detected rearrangement in *PTEN* involved reciprocal translocation with *PDE1C*. RNAsum identified a *PTEN::PDE1C* fusion (*PTEN* exon 2 - *PDE1C* exon 17), which caused a frameshift and predicted an aberrant PTEN protein truncated early in the phosphatase domain. Coupled with its low expression (2^nd^ percentile), these findings strongly supported the loss of function of *PTEN* (Fig. 4B).

**Fig. 4.**
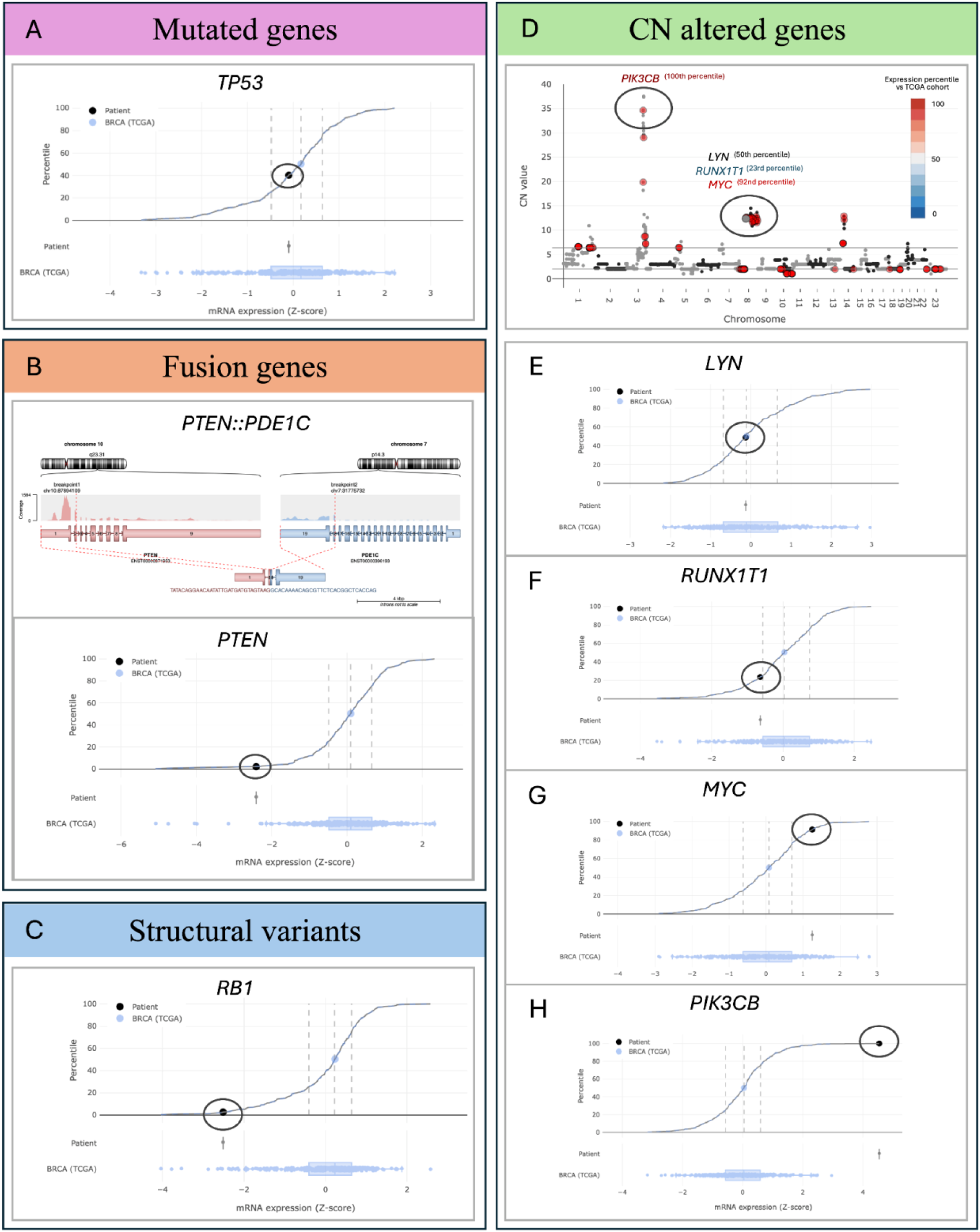
RNAsum visualisations supporting the key genomic and transcriptomic findings from the analysis of TNBC patient. Panel **A** shows the *TP53* expression level at the 41^st^ percentile relative to the TCGA breast cancer cohort. Panel **B** highlights the *PTEN*::*PDE1C* fusion transcript detected by WTS, indicating that an aberrant PTEN protein was truncated early in the *PTEN* phosphatase domain. This fusion is coupled with low *PTEN* expression (2^nd^ percentile), which is consistent with a loss of *PTEN* function. Panel **C** depicts the low expression of *RB1* (2^nd^ percentile compared with TCGA) supporting frameshift mutation and loss of function following tandem duplication involving exon 15, as confirmed by WTS. Panel **D** displays the amplification of chromosome 8q, including 32 cancer-related genes, as well as the chr3q22 region encompassing *PIK3CB*. *PIK3CB* and three genes located within the chromosome 8q amplification, including *MYC*, *LYN* and *RUNX1T1*, are colour-encoded by their expression percentile rank relative to the TCGA breast cancer cohort (blue: low expression; grey: median; red: high expression). Panels **E**-**G** show the expression levels of other genes within the 8q amplification, including *LYN* and *RUNX1T1*, which were not highly expressed, and *MYC*, which was highly expressed (92^nd^ percentile). Panel **H** illustrates the amplification of *PIK3CB* in the chr3q22 region, with expression at the 100^th^ percentile relative to the TCGA cohort. *PIK3CB* overexpression is noted as a potential driver of PI3K signalling but remains classified as a variant of uncertain significance.

The WGS-detected rearrangement in *RB1* involved a tandem duplication of exon 15. WTS confirmed the expression of transcripts carrying the duplication, resulting in frameshift and predicting truncation after Phe473, which is the N-terminus of the essential RB1 pocket domain. RNAsum also reported low *RB1* expression (Fig. 4C) in the 2^nd^ percentile compared with the TCGA Breast Cancer cohort, adding confidence to the DNA-based interpretation. WGS also revealed amplification of the entire long arm of chromosome 8 (CN range between 10.8 and 14.5). This large amplicon contained 32 cancer-related genes (Fig. 4D), 3 of which are known oncogenes according to the OncoKB [23,24] database: *LYN* (Fig. 4E), *RUNX1T1* (Fig. 4F) and *MYC* (Fig. 4G). RNAsum identified only MYC as highly expressed (92^nd^ percentile relative to the TCGA Breast Cancer cohort). *MYC* amplification and expression were reported as potential biomarkers for inclusion in a clinical trial (Australian and New Zealand Clinical Trial Registry ACTRN12620001146987).

In addition, amplification of the chr3q22 region, encompassing *PIK3CB* (CN of 34.6), was detected. *PIK3CB* encodes p110β, an activating subunit of PI3 kinase (PI3K). In contrast to its paralogue *PIK3CA*, mutations of *PIK3CB* are rarely observed in TNBC [37–39]. RNAsum reported high *PIK3CB* expression (100^th^ percentile when compared with the TCGA breast cancer cohort) Fig. 4H. This amplification was reported as a variant of unknown clinical importance due to the ability of p110β to drive PI3K signalling [40].

#### Case study #2

We performed whole-genome and transcriptome sequencing on a fresh biopsy sample from a woman in her late 20’s with hormone receptor-positive metastatic breast cancer who had progressed on aromatase inhibitor and gonadotropin-releasing hormone agonist therapy (Table S2, Case 43).

Amplification of *CCND1* (12 copies) and *FGFR1* (20 copies) was detected by WGS (Fig. 5A) and RNAsum reported high expression of both genes relative to the TCGA Breast Cancer cohort (100th and 98th percentiles, respectively) (Fig. 5B). These two genes are recurrently co-amplified in hormone receptor-positive breast cancer [41].

**Fig. 5.**
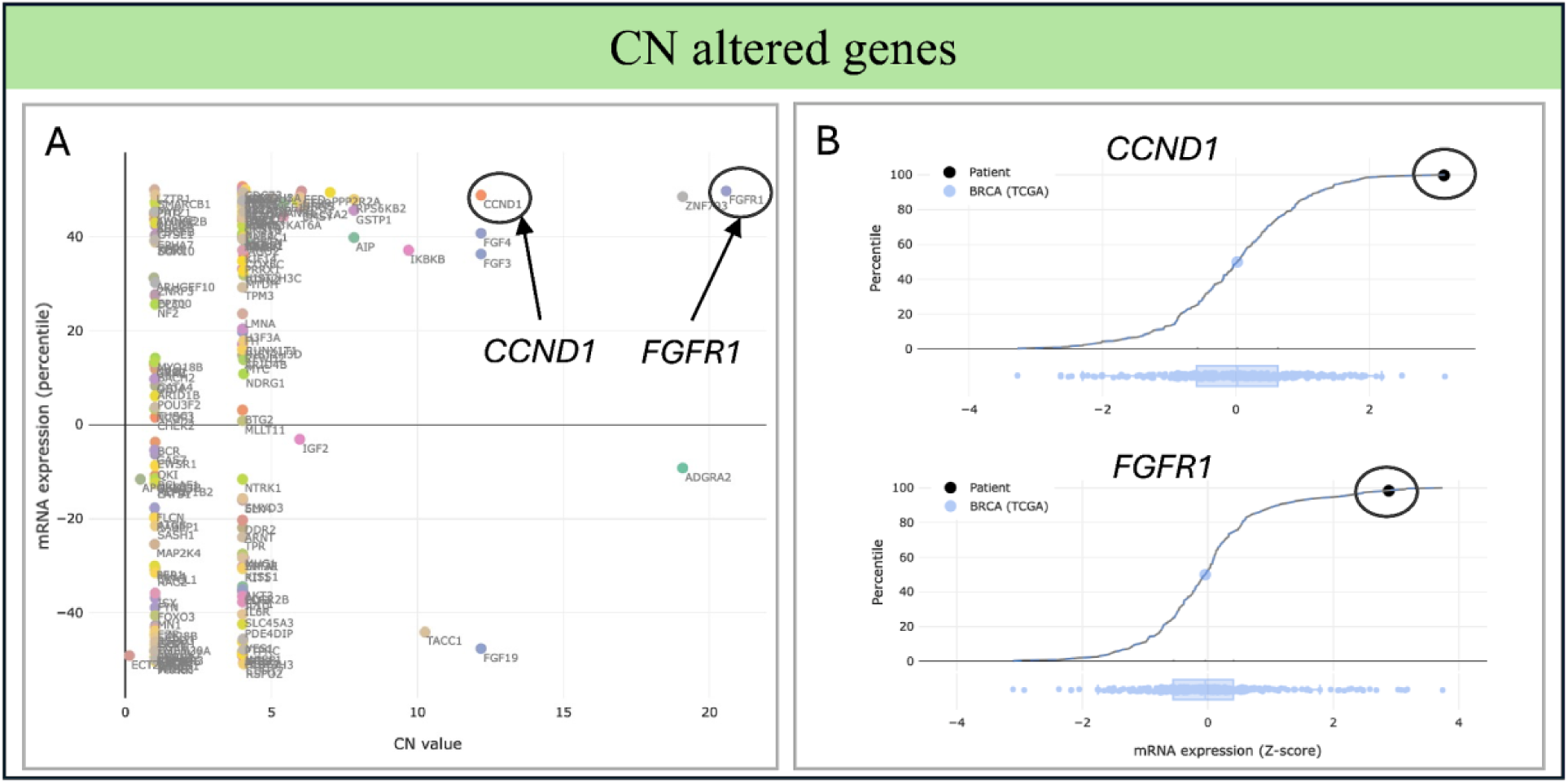
RNAsum analysis and visualisation of *CCND1* and *FGFR1* co-amplification in metastatic breast cancer. Panel **A** presents a scatter plot of CNVs detected by WGS, plotted against corresponding gene expression levels from WTS. The recurrently co-amplified genes *CCND1* (12 copies) and *FGFR1* (20 copies) are highlighted, where both show markedly elevated expression. Panel **B** shows plots illustrating *CCND1* and *FGFR1* expression levels in the patient’s tumour compared with TCGA, positioned at the 100^th^ and 98^th^ percentiles, respectively.

In addition, a hotspot missense mutation (c.1613A>G p.D538G) and complex structural rearrangement of ESR1, encoding oestrogen receptor alpha (ERα), were detected. Genetic alterations in ESR1 are commonly detected in hormone receptor-positive breast cancer patients with acquired resistance to prolonged endocrine therapy [42–47]. WGS identified interchromosomal translocations involving *ESR1* and *PLEKHG1* on chromosome 6, and *HNRNPM* on chromosome 19 (Fig. 6A). RNAsum reported robust expression of an in-frame fusion transcript coupling *ESR1* exon 4 to *PLEKHG1* exon 12, clarifying the interpretation of this complex rearrangement (Fig. 6B-D). The p.D538G substitution observed in this patient confers resistance to oestrogen deprivation therapies but predicts sensitivity to ER degraders such as elacestrant. On the other hand, few clinical studies suggest that fusion genes involving *ESR1* exons 1-6, which encode ERα proteins that lack binding sites for selective ER modulators and degraders, may predict resistance to ER degrader therapy [45,46], highlighting the complexity of acquired endocrine therapy resistance mechanisms. Thus, elucidation of a complex *ESR1* rearrangement in this case illustrates the potential of RNAsum in guiding clinical management.

**Fig. 6.**
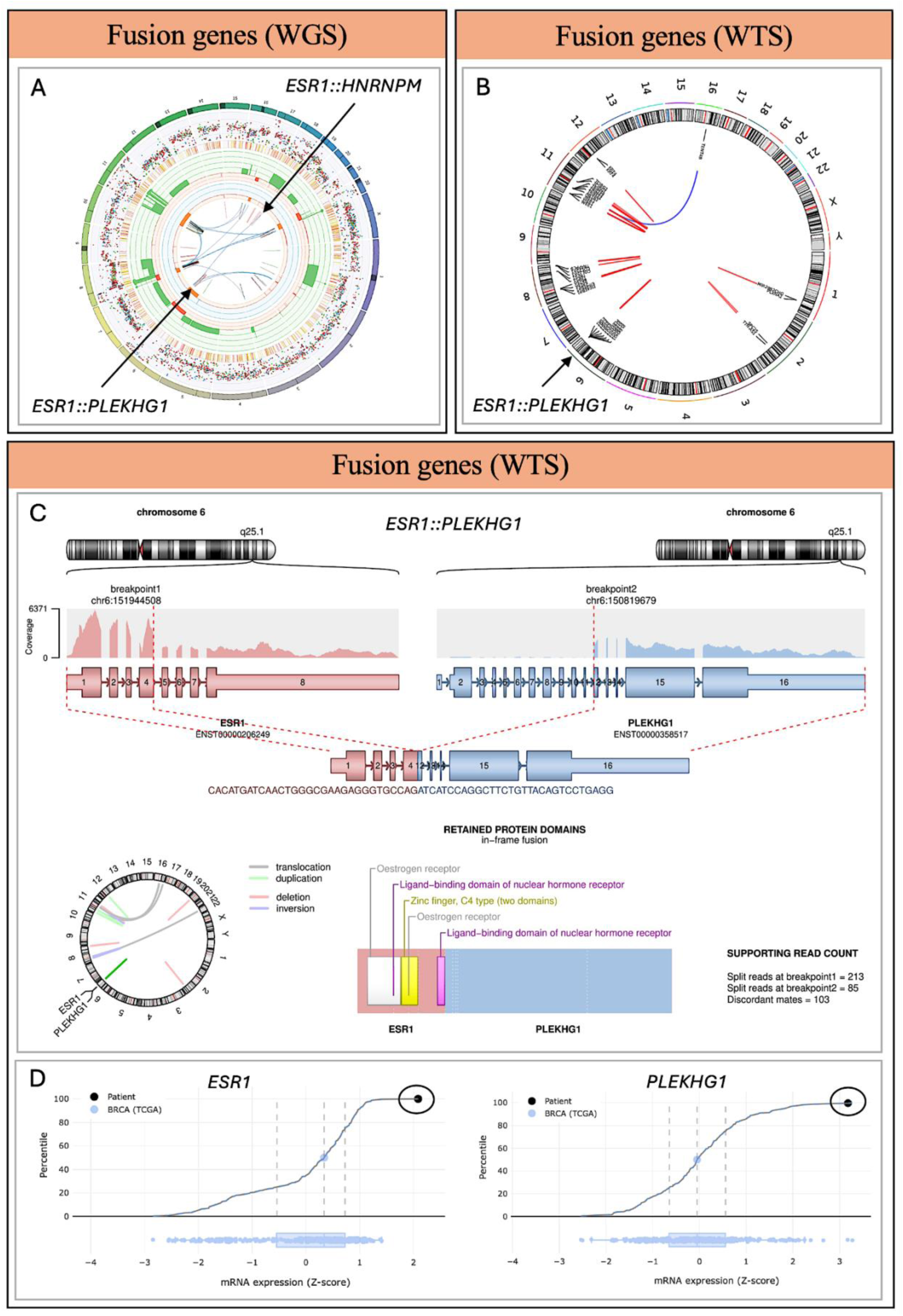
RNAsum visualisation of a complex *ESR1* structural rearrangement detected in hormone receptor–positive metastatic breast cancer. Panels **A**-**D** summarise genomic and transcriptomic evidence from WGS and WTS. **A** Circos plot based on WGS data showing genome-wide somatic alterations. Outer rings display chromosomes (dark regions = centromeres/heterochromatin), somatic variants (SNPs coloured according to the type of base change, e.g. C>T/G>A in red, and Indels in yellow/red), CN changes (red = losses, green = gains), and minor allele CN (orange = LOH, blue = gain). Inner arcs represent structural variants (blue = translocations, red = deletions, green = duplications, black = inversions). WGS revealed multiple *ESR1* rearrangements, including *ESR1*::*HNRNPM* (chr19) and *ESR1*::*PLEKHG1* (chr6), indicating a complex event not fully resolved by WGS alone. **B** WTS Circos plot showing high-confidence fusion transcripts (red lines), including *ESR1*::*PLEKHG1*, which clarified the functional consequence of the complex WGS rearrangement. **C** Schematic of the *ESR1*::*PLEKHG1* fusion detected by WTS, joining *ESR1* exon 4 with *PLEKHG1* exon 12. The in-frame fusion lacks key ER modulator and degrader binding sites. RNA read support and domain structures are shown. WTS confirmed this as the expressed and functional rearrangement, distinguishing it from the additional *ESR1*::*HNRNPM* event. **D** Expression profiles for *ESR1* and *PLEKHG1*, both at the 100^th^ percentile relative to the TCGA cohort, confirming robust transcription of the fusion partners and supporting the fusion’s functional and clinical relevance.

## Discussion

We present RNAsum, a robust open-source bioinformatics reporting tool designed to address the complex challenges of integrating and summarizing genomic and transcriptomic data in cancer research. RNAsum leverages RNA-seq data from cancer patients to either verify and complement whole-genome profiling results or provide comprehensive summary statistics from WTS data. This tool tackles several key obstacles in the field, including the tumour-only nature of transcriptome sequencing, the need to harmonize and interpret outputs from diverse analyses and the lack of cancer-specific reference data and complex integration tools. By offering a simple unified approach to data interpretation, RNAsum aims to enhance our understanding of cancer biology and support informed clinical decision-making in precision oncology.

RNAsum incorporates transcriptome profiling into a precision oncology framework by utilizing landmark TCGA RNA-seq data, thereby increasing confidence in the clinical utility of findings. RNAsum integrates data at multiple levels by combining mRNA profiles from the patient and external reference cohorts, overlaying expression data with genomic changes, and annotating findings with clinically relevant information from public knowledge databases. This comprehensive integration process enables the identification and interpretation of cancer-relevant molecular aberrations, facilitating precision oncology by streamlining genomics research, accelerating discoveries, and translating them into actionable clinical insights. Based on comprehensive benchmarking (Fig. S1), we recommend using the representative TCGA reference cohort mode for routine workflows, particularly when dealing with memory-demanding pan-cancer analyses. This mode offers improved scalability with minimal loss of precision, as 99% of known cancer genes show less than a 10^th^ percentile point difference compared with the full mode, with discrepancies primarily affecting low-expression genes.

RNAsum generates an interactive HTML-based report summarizing clinically relevant information, including mutated genes, fusion genes, SVs, CN alterations, as well as customizable gene sets (e.g. immune markers and DNA damage repair genes). The report provides comprehensive summaries of mRNA expression levels, prioritised findings based on whole-genome sequencing results and public databases, and visual data summaries for quality control and interpretation. Despite the growing adoption of WGTS in precision oncology, dedicated tools for automated single-patient transcriptomic interpretation remain limited. Existing approaches either require substantial infrastructure, including production-ready database instances and continuous management overhead [48] or are primarily designed for cohort-level exploratory analysis rather than individual patient reporting [49], or perform RNA outlier detection independently of patient-specific WGS findings [50]. RNAsum addresses a distinct gap as an automated, report-centric tool that directly integrates user-provided WGS and WTS data from an individual patient with matched tumour-type reference cohort, without requiring dedicated database infrastructure. RNAsum contextualises WGS-detected variants through their transcriptional consequences, quantifying relative expression levels, detecting aberrant transcripts and fusion events, and anchoring findings to a clinically relevant reference population. In doing so, it translates complex multi-omic outputs into self-contained, interactive HTML report suitable for direct use in precision oncology workflows and molecular tumour board discussions. As a platform-agnostic R package that can be installed and run with minimal technical expertise or management overhead, it provides a complementary and flexible alternative to infrastructure-heavy reporting solutions, making automated WGTS interpretation accessible within existing clinical bioinformatics pipelines.

Our evaluation of RNAsum’s clinical utility highlights its robust performance in supporting genomic findings at the transcriptomic level. In a comprehensive review of 60 cases with matched WGS and WTS data, RNAsum provided additional supporting information in 68% of the clinically reportable variants where transcriptomic confirmation was deemed possible. Notably, in 87% of cases, at least half of the WGS-detected variants were supported by WTS data, with full support for all possible variants observed in 52% of cases. This high concordance across various mutation types underscores the effectiveness of RNAsum in enhancing the accuracy and reliability of clinical genomic interpretations, supporting its integration into precision oncology workflows and potentially improving patient management and health outcomes.

While traditional molecular pathology techniques such as FISH and IHC are widely used for identifying genomic changes, the integration of RNA-seq data with genomic analysis has proven invaluable for detecting and characterising complex genomic rearrangements, particularly gene fusions. This multiomic approach enables the identification of in-frame chimeric transcripts and the assessment of their functional consequences, which may not be apparent from genomic data alone. Our method has demonstrated the significant utility of WGTS analysis in refining cancer diagnoses and facilitating the discovery of potential therapeutic targets. Using RNAsum, we successfully characterised a complex *HERPUD1*::*RAF1* fusion gene in a case of pancreatic acinar carcinoma [51] and a diagnostic *CIC*::*DUX4* fusion in a case of *CIC*-rearranged sarcoma [52]. WGTS analysis was instrumental in determining diagnoses for 32% of sarcoma patients in an adolescent and young adult cohort [53], and for 66% of patients in a Cancer of Unknown Primary (CUP) cohort [54]. Finally, RNAsum supported the corroboration of a novel *RET*::*SEPTIN9* fusion in a pheochromocytoma patient, leading to successful treatment with an *RET* inhibitor [55].

RNAsum offers great flexibility in data processing and parameter selection, but it requires users to provide specific WGTS analysis inputs in a compatible format for integration into the reporting. This presents opportunities for developing modules that can support outputs from various bioinformatics workflows and tools. Although TCGA provides a rich resource of cancer-specific datasets for gene expression comparisons, it has been shown to exhibit unwanted variation arising from library size, tumour purity, and batch effects [56]. Hence, the results should be interpreted within a specific biological context and corroborated through orthogonal evidence or supported by the literature.

To ensure efficient data and dependency management, the RNAsum codebase was developed as an R-package that can be easily accessed and downloaded from GitHub. It offers flexibility in execution, as it can be run on either an HPC or a cloud infrastructure, and is also available as a docker container. The TCGA reference data utilized by RNAsum are conveniently managed through an R data package [57], which is also accessible via GitHub. Moreover, RNAsum is a cost-effective solution that can be seamlessly adapted to run on outputs from WTS analysis exclusively or from both WGS and WTS analysis.

## Conclusion

We developed RNAsum, a novel bioinformatics method that deploys a comprehensive approach to data preprocessing, quantification, and integration of multiomics datasets. RNAsum represents a significant advancement in RNA-seq data analysis, seamlessly integrating WGS and WTS data to provide critical insights for cancer patient genomes. Additionally, we demonstrate that by successfully achieving its aims of delivering clinically relevant information and prioritising results for therapeutic intervention for clinical cases, RNAsum enhances patient prognosis, management, and health outcomes in precision oncology settings. The tool’s potential to improve data interpretation and accelerate discoveries in genomics research is substantial, paving the way for streamlining research efforts and driving personalised medicine initiatives. To enable its wider applicability and usage, RNAsum is available as an R package that can be downloaded from GitHub https://github.com/umccr/RNAsum/tags.

## Data Availability

The Cancer Genome Atlas (TCGA) data in the present study are available online at the National Cancer Institute (NCI) Genomic Data Commons (GDC) Data Portal (https://portal.gdc.cancer.gov/). The clinical case data presented in this study are available upon request from the corresponding author and pending approval from the VCCC PRECISION program investigators. The data are not publicly available due to ethics restrictions.

## Availability and requirements

Project name: RNAsum

Project home page: https://github.com/umccr/RNAsum

Operating system(s): Platform independent

Programming language: R

Other requirements: R 4.0.0 or higher Licence: MIT

## List of abbreviations

WGS: whole-genome sequencing
WTS: whole-transcriptome sequencing
TCGA: The Cancer Genome Atlas
HTS: High-throughput sequencing
SNV: single nucleotide variant
Indels: small insertions and deletions
SV: structural variant
CNV: copy-number variant
TMB: tumour mutational burden
HRD: homologous recombination deficiency
MSI: microsatellite instability
WGTS: whole-genome and transcriptome sequencing
RNA-seq: RNA sequencing
NCI: National Cancer Institute
GDC: Genomic Data Commons
API: Application Programming Interface
GOIs: genes of interest
CGI: Cancer Genome Interpreter
CIViC: Clinical Interpretation of Variants in Cancer
VICC: Variant Interpretation for Cancer Consortium
CPM: counts per million
PCGR: Personal Cancer Genome Reporter
PCA: principal component analysis
RLE: relative log expression
TNBC: triple-negative breast cancer
LOH: loss of heterozygosity
ERα: oestrogen receptor alpha
CUP: Cancer of Unknown Primary

## Declarations

### Ethics approval and consent to participate

The VCCC PRECISION program received ethics approval from the Peter MacCallum Cancer Centre Human Research Ethics Committee (HREC/48455/PMCC-2018).

### Consent for publication

Informed consent was obtained from all subjects involved in the study.

### Availability of data and materials

RNAsum is available as an R package distributed under the MIT Licence (https://opensource.org/licenses/MIT), allowing unrestricted reuse with appropriate attribution, through GitHub (https://github.com/umccr/RNAsum) and Anaconda (https://anaconda.org/umccr/r-rnasum) repositories.

The original TCGA [19] datasets used as reference cohorts are available from the NCI Genomic GDC Data Portal (https://portal.gdc.cancer.gov/) via the GDC API (https://gdc.cancer.gov/developers/gdc-application-programming-interface-api) and data release 40.0 (March 29, 2024, https://docs.gdc.cancer.gov/Data/Release_Notes/Data_Release_Notes/#data-release-400). The clinical case data presented in this study are available upon request from the corresponding author and pending approval from the VCCC PRECISION program investigators. The data are not publicly available due to ethics restrictions.

RNAsum reports from these case studies are deposited in the Zenodo repository (DOI: 10.5281/zenodo.17353510).

### Author Contributions

SMG, OH, SK and JM conceptualised the study. SK and JM implemented the software. KP was responsible for sequencing. LT, LNV and JHAV contributed to data curation and interpretation. SK and JM drafted the manuscript. JHAV and LNV proposed and wrote the case studies. SK and JM prepared the figures and data summaries. SMG and SJL acquired funding and ethics approval. All authors reviewed and approved the case study.

## Acknowledgements

We would like to acknowledge the Collaborative Centre for Genomics Cancer Medicine and Genomics Platform for WGTS and bioinformatics analysis. We would also like to thank all patients, the clinicians and the pathologists who facilitated data acquisition and generation.

## Supplementary Material

Provided in a separate file “RNAsum_suppl”

