## Supplemental figures and table S1 and S3 for "RNAsum: a tool for personalised genome and transcriptome interpretation for improved cancer diagnostics"

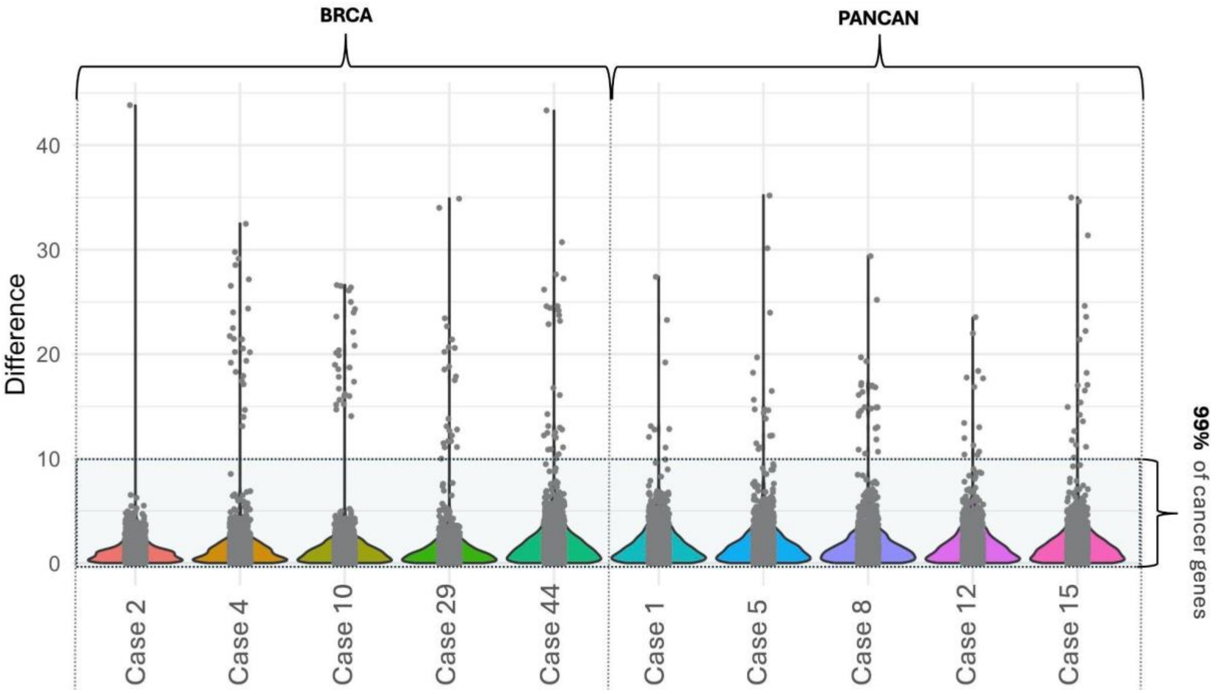


Fig. S1 Benchmarking Cancer Gene Percentile using “full” or “representative” reference cohorts. Violin plot presenting discordant cancer genes, which on average make up 1% of all cancer genes in investigated cases, observed when comparing results from “full” or “representative” external reference cohort mode (see Table S1). Benchmarking was performed using five cases combined with tumour-matched and five cases combined with pan-cancer TCGA cohorts. The y-axis represents differences in percentile values relative to the corresponding reference cohort.

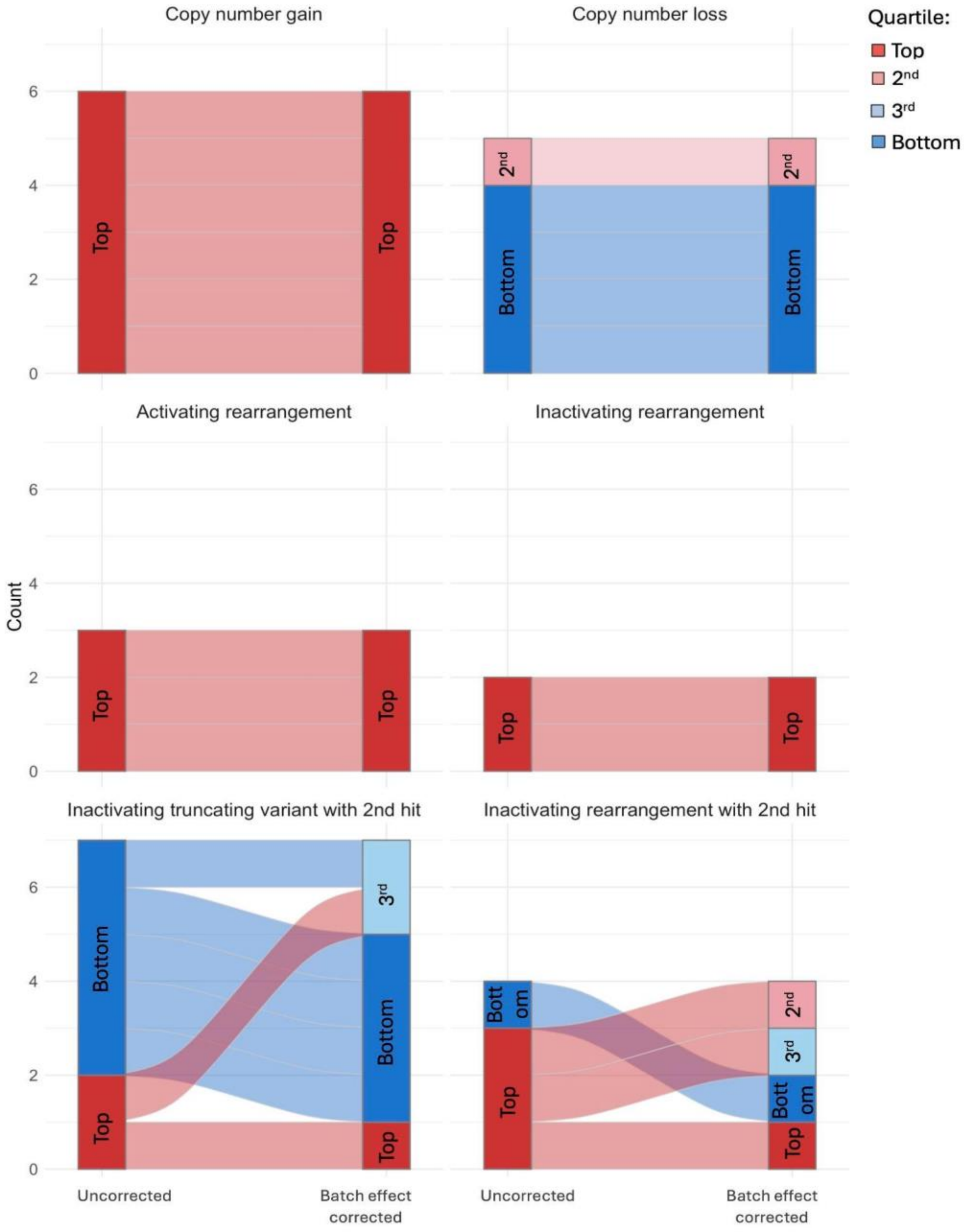

Copy number gain
Copy number loss
Activating rearrangement
Inactivating rearrangement
Inactivating truncating variant with 2nd hit
Inactivating rearrangement with 2nd hit
Count
0
2
4
6
Top
2nd
3rd
Bottom
Bott om
Uncorrected
Batch effect corrected
Quartile:
Top
2nd
3rd
Bottom

Fig. S2 Impact of batch-effect correction on expression percentiles. Alluvial plots illustrating the distribution of expression percentile categories defined in WTS from 10 patient cases (five combined with tumour-matched and five with pan-cancer external reference cohorts) for genes with clinically relevant variant types detected by WGS, before (uncorrected) and after batch-effect correction (batch effect corrected). Each ribbon represents the flow of variants between quartile categories, with colours denoting relative expression levels (blue: lower quartiles, red: higher quartiles). The plots demonstrate how batch-effect correction can alter the categorisation of variants across expression percentiles, thereby impacting interpretation of expression changes linked to specific variant types. Across all 27 clinically relevant variants, only two inactivating truncating variants with a second hit and three inactivating rearrangements with a second hit fell into different quartile categories, indicating that batch-effect correction using the internal reference cohort has little impact on the expression levels of genes affected by clinically relevant genomic alterations.

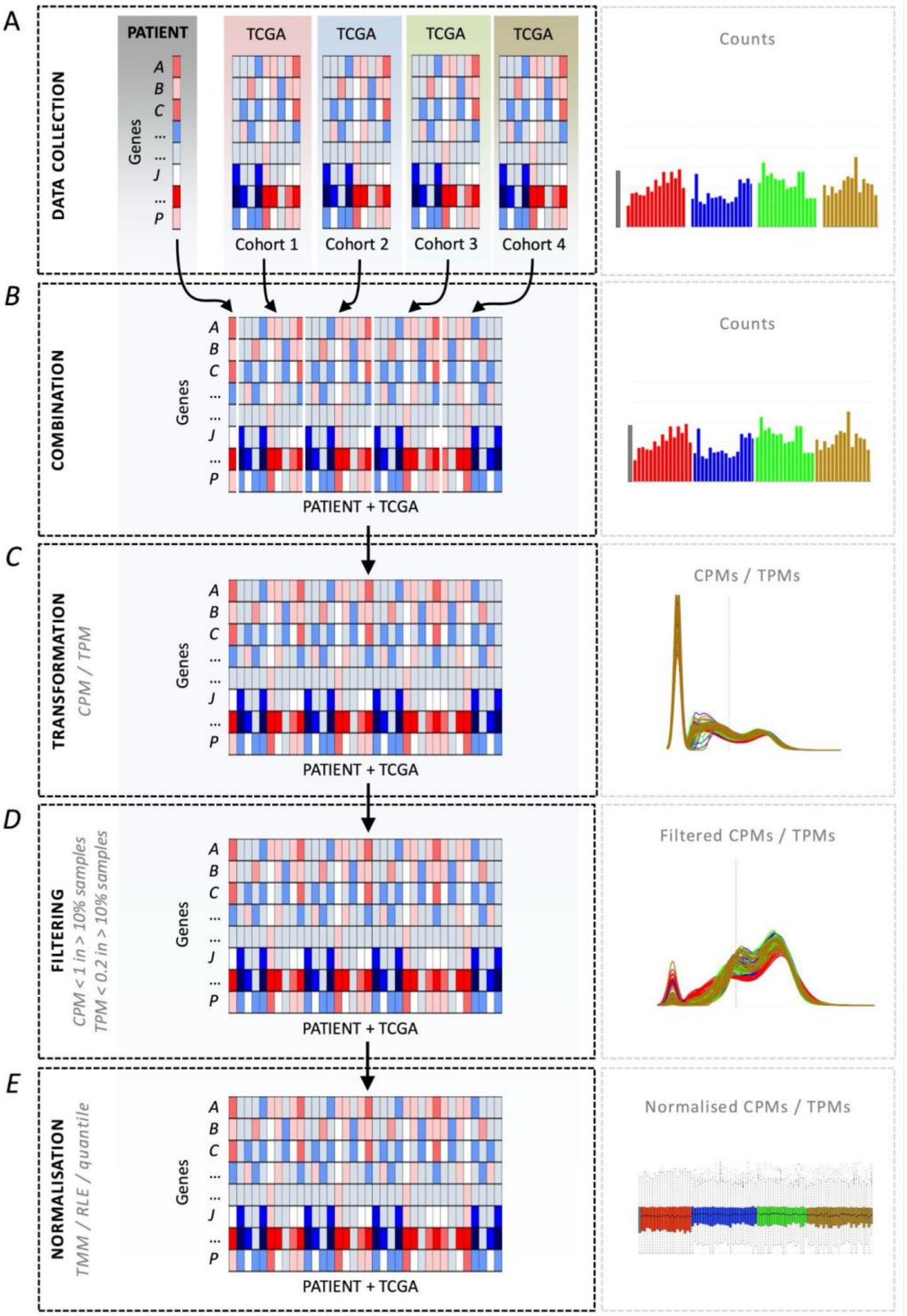


Fig. S3 Read counts processing scheme. **A** Read count data from the following two datasets are used as input: (1) patient sample and (2) external reference cohort from TCGA. **B** Data combination following subsetting patient sample and TCGA reference datasets to include common genes. **C** Data transformation performed by converting counts to CPM (Counts Per Million; default) or TPM (Transcripts Per Kilobase Million) values. **D** Filtering out genes with low counts (CPM or TPM < 1 in more than 90% of samples). **E** Data normalization to account for sample-specific effects.

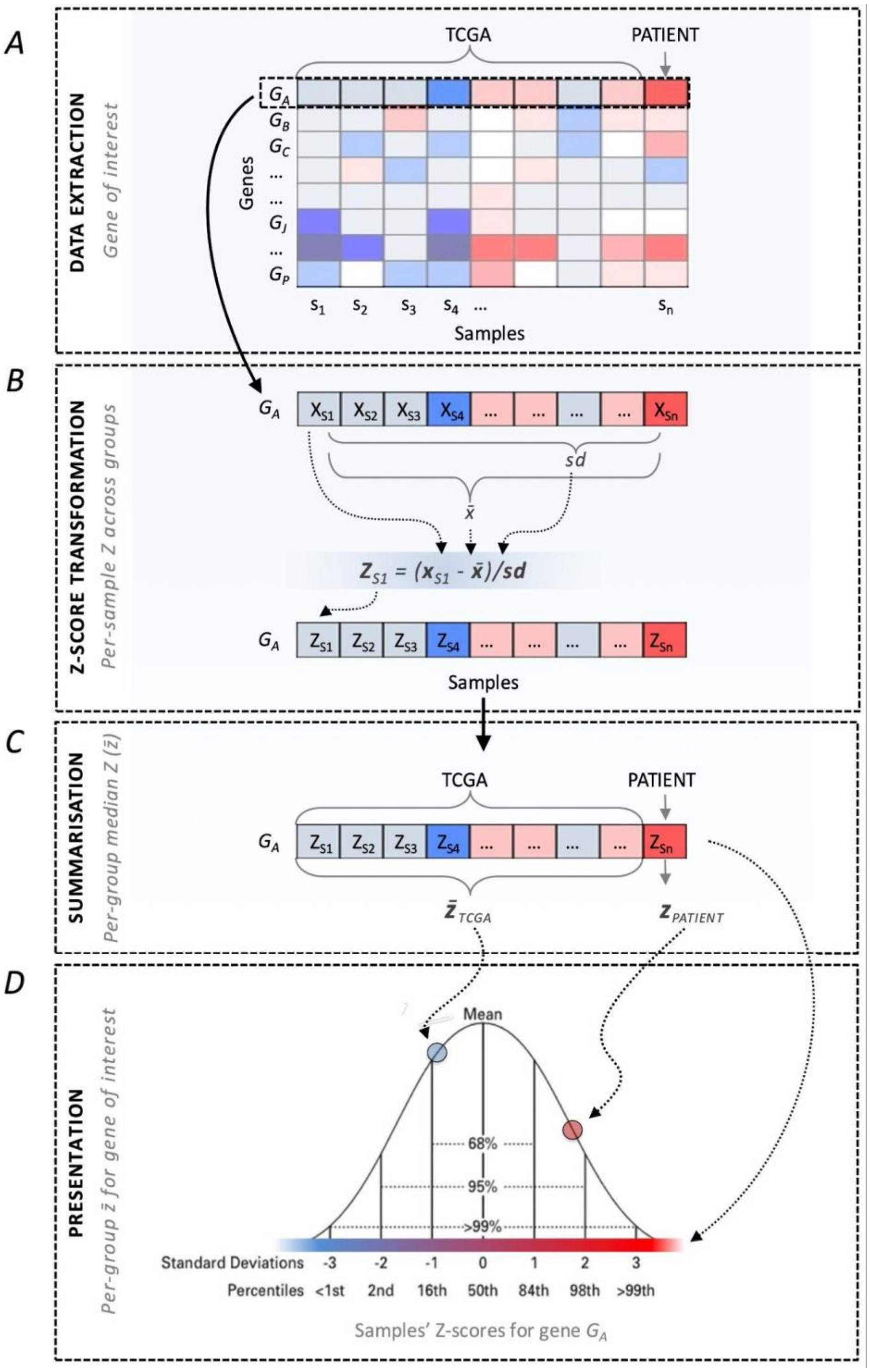


Fig. S4 Z-transformation (standardization) scheme. **A** Extract expression values across all samples for a given gene. **B** Compute Z-scores for individual samples for that gene. **C** Compute median Z-scores for external reference dataset. **D** Present patient sample Z-score in the context of the reference cohorts' median Z-score.

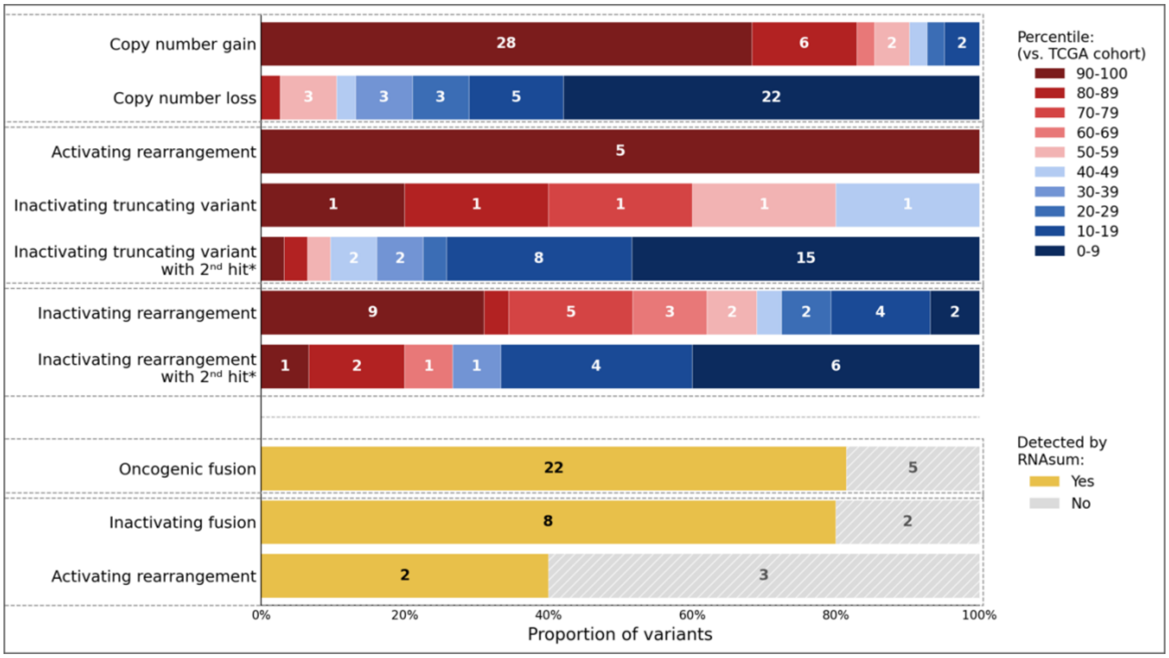


Fig. S5 Transcriptomic corroboration of clinically relevant genomic alterations through RNAsum at decile resolution. The bar plot summarises the genomic alterations reported across 60 cases, categorised by WTS-based evidence, displayed using 10-percentile expression bins relative to the matched TCGA reference cohort. The stacked segments within each bar indicate the type of WTS support: relative expression deciles for copy number alterations, activating and inactivating variants (with or without a second hit*), and detection of aberrant transcripts for structural rearrangements. The numerical labels within the bars indicate the number of variants supported by each WTS evidence category. This overview highlights the value of integrating WGS and WTS data to corroborate clinically reportable genomic variants, demonstrating RNAsum's role in contextualising expression changes and fusion events alongside genomic alterations.

**Includes inactivating variants on the X chromosome in males.*

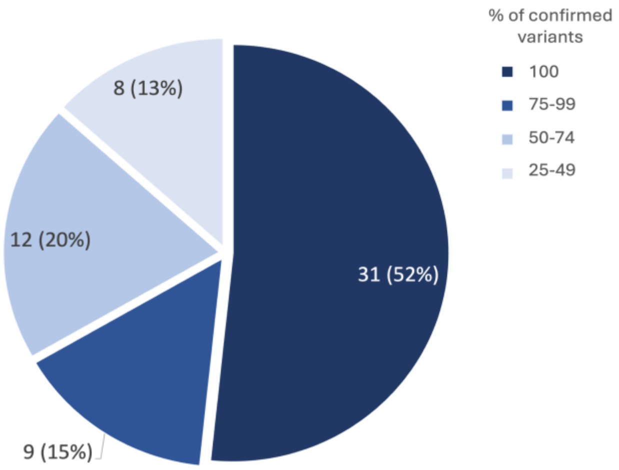


Fig. S6 Pie chart showing the proportion of clinically reportable variants confirmed by WTS across cases. Each slice represents the percentage of confirmed variants within a specific category, with the number inside each slice indicating the count of cases in that category.

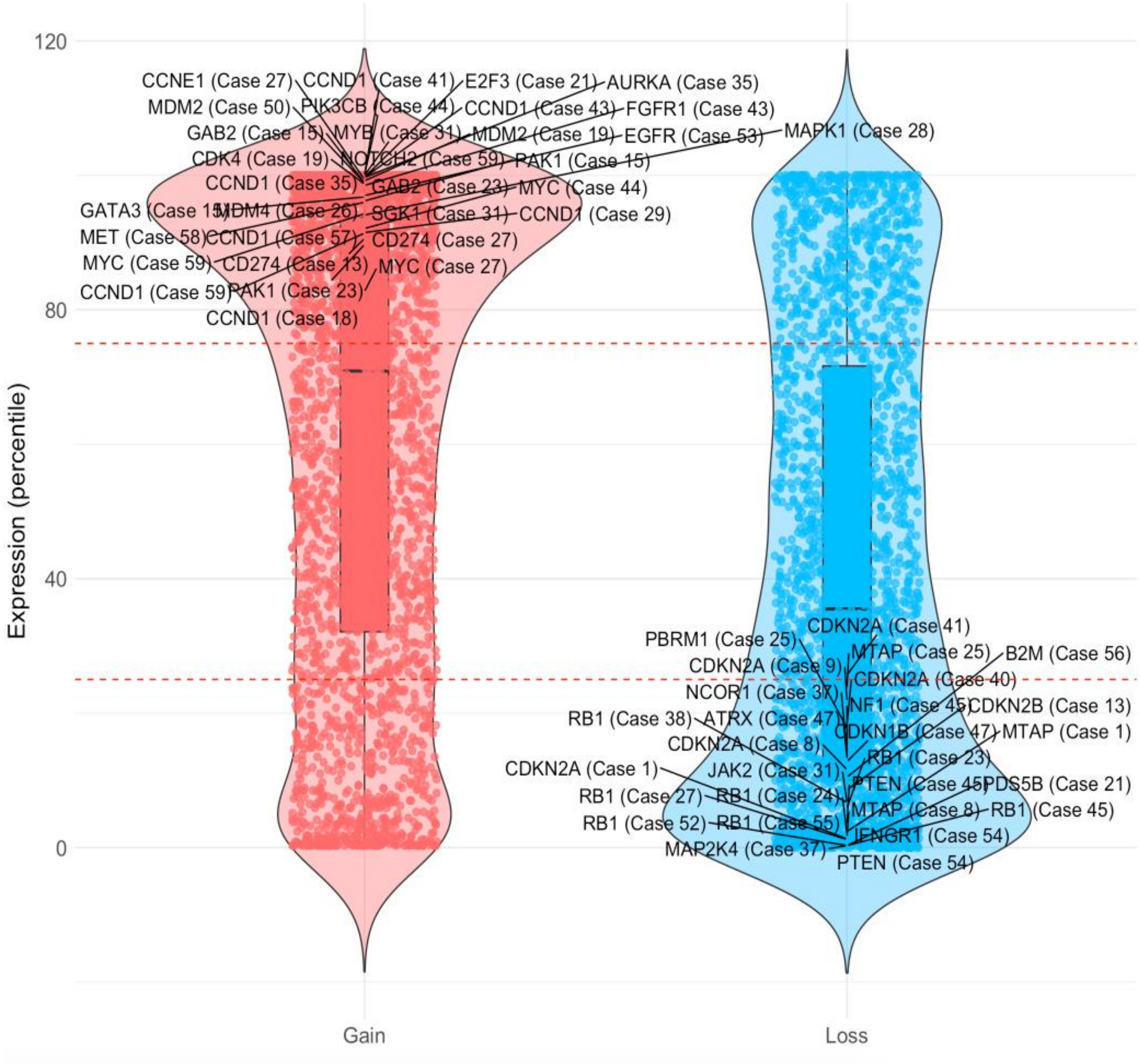


Fig. S7 Expression profiles of clinically relevant copy number variants. Violin and box plots showing expression percentiles for CN gain and loss variants identified by WGS across 60 patient cases and included in the RNAsum clinical applicability evaluation. Labelled genes (with case numbers in parentheses) represent clinically reportable variants supported by RNA-level evidence from RNAsum, where support was defined as a substantial expression increase for CN gains (top quartile relative to the TCGA reference cohort; upper red dashed line) or a substantial expression decrease for CN losses (bottom quartile; lower red dashed line). Remaining points correspond to the rest of CN-altered genes detected by WGS.

Supplementary Note 1: TCGA strandedness characterisation.

To characterise the strandedness of the TCGA reference data used by RNAsum, we performed a pan-cancer census of all 11,505 TCGA samples available through the NCI Genomic Data Commons (GDC, Data Release 40.0), examining STAR alignment summary statistics, specifically the N_noFeature ratios under stranded-first and stranded-second quantification modes, where a ratio close to unity between the two modes is diagnostic of non-strand-specific library preparation. This analysis confirmed that 32 of 33 TCGA projects (96.6%; n = 11,114 samples) are genuinely unstranded, consistent with historical non-strand-specific poly(A) library preparation across the majority of TCGA cancer types. Only TCGA-GBM constitutes a mixed cohort (n = 391 samples, 3.4% of the total). This is in line with the GDC's own harmonisation policy, which explicitly states that all TCGA RNA-Seq reads are treated as unstranded during analyses. RNAsum accordingly uses unstranded GDC counts for all reference cohort expression metrics. To quantify the potential impact on percentile rankings when a patient sample is prepared with a strand-specific protocol (e.g., dUTP/ISR, increasingly common in clinical workflows), we performed a structural analysis using GENCODE v36, identifying protein-coding genes with ≥1,000 bp antisense overlap with a protein-coding or lncRNA partner. The 1,000 bp threshold was chosen as the minimum at which antisense co-occupancy encompasses at least one full exon-equivalent of genomic sequence, below which per-nucleotide read contamination is negligible. This yielded 6,782 protein-coding genes (34.0%), including 51 clinically actionable cancer genes. Of these, 21 genes carry antisense overlaps ≥10,000 bp, approximately one-third or more of a typical gene body, and the point at which antisense co-occupancy is large enough to produce clinically non-trivial percentile displacement even for moderately expressed genes. These 21 genes (*ALK*, *BRCA1*, *CDKN2A*, *NF1*, *RB1*, *RUNX1*, *ATM*, *KMT2A*, *BCL6*, *FBXW7*, *MSH2*, *MSH6*, *PDCD1LG2*, *FANCD2*, *KDR*, *CDK6*, *ERBB2*, *ESR1*, *EGFR*, *CTNNB1*, and *SMARCA4*) are listed in the RNAsum HTML report, which flags them with a recommendation for additional interpretive caution when the patient sample is identified as strand-specific.

Table S1 Table summarising the 33 TCGA cancer types (10,079 cancer patients in total) obtained from NCI GDC Data Portal (data release 40.0, March 29, 2024) and used in RNAsum workflow as reference cohorts. The patient's number included in the individual cohorts using either “representative” (default) or “full” reference cohort modes are indicated in the last two columns. *Tissue: 1 - solid tissue; 3 - peripheral blood*

| Reference cohort | Cancer type | Tissue | Patients number (representative cohort) | Patients number (full cohort) |
|---|---|---|---|---|
| PANCAN | All TCGA cancer types (pan-cancer) | 1,3 | 330 | 10,079 |
| BRCA | Breast Invasive Carcinoma | 1 | 300 | 1,102 |
| STAD | Stomach Adenocarcinoma | 1 | 300 | 369 |
| LUAD | Lung Adenocarcinoma | 1 | 300 | 349 |
| LIHC | Liver Hepatocellular Carcinoma | 1 | 300 | 345 |
| PRAD | Prostate Adenocarcinoma | 1 | 300 | 336 |
| KIRC | Kidney Renal Clear Cell Carcinoma | 1 | 300 | 335 |
| THCA | Thyroid Carcinoma | 1 | 300 | 332 |
| LUSC | Lung Squamous Cell Carcinoma | 1 | 300 | 332 |
| HNSC | Head and Neck Squamous Cell Carcinoma | 1 | 300 | 330 |
| LGG | Brain Lower Grade Glioma | 1 | 300 | 309 |
| COAD | Colon Adenocarcinoma | 1 | 257 | 257 |
| KIRP | Kidney Renal Papillary Cell Carcinoma | 1 | 252 | 252 |

| | | | | |
|---|---|---|---|---|
| BLCA | Bladder Urothelial Carcinoma | 1 | 246 | 246 |
| OV | Ovarian Serous Cystadenocarcinoma | 1 | 220 | 220 |
| SARC | Sarcoma | 1 | 214 | 214 |
| PCPG | Pheochromocytoma and Paraganglioma | 1 | 177 | 177 |
| CESC | Cervical Squamous Cell Carcinoma and Endocervical Adenocarcinoma | 1 | 171 | 171 |
| UCEC | Uterine Corpus Endometrial Carcinoma | 1 | 168 | 168 |
| PAAD | Pancreatic Adenocarcinoma | 1 | 150 | 150 |
| TGCT | Testicular Germ Cell Tumours | 1 | 149 | 149 |
| LAML | Acute Myeloid Leukaemia | 3 | 145 | 145 |
| ESCA | Esophageal Carcinoma | 1 | 142 | 142 |
| GBM | Glioblastoma Multiforme | 1 | 141 | 141 |
| THYM | Thymoma | 1 | 118 | 118 |
| SKCM | Skin Cutaneous Melanoma | 1 | 100 | 100 |
| READ | Rectum Adenocarcinoma | 1 | 87 | 87 |
| UVM | Uveal Melanoma | 1 | 80 | 80 |
| ACC | Adrenocortical Carcinoma | 1 | 78 | 78 |
| MESO | Mesothelioma | 1 | 77 | 77 |

| | | | | |
|---|---|---|---|---|
| KICH | Kidney Chromophobe | 1 | 59 | 59 |
| UCS | Uterine Carcinosarcoma | 1 | 56 | 56 |
| DLBC | Lymphoid Neoplasm Diffuse Large B-cell Lymphoma | 1 | 47 | 47 |
| CHOL | Cholangiocarcinoma | 1 | 34 | 34 |

Table S2 Table presenting an overview of clinically reportable genomic alterations detected by WGS across the 60 reviewed cases. For each case, it lists the cancer type, affected gene, and the type of the genomic variant. The table indicates whether individual genomic alterations are supported by data from WTS, including increased expression, loss of expression, aberrant splicing or fusion events. Cases 43 and 44 are described in detail as Case study #2 and Case study #1, respectively, in the main text.

Provided as a separate excel file "Table_S2.xlsx".

Table S3. Clinical cases where RNAsum critically informed variant interpretation. Case 43 is described in detail as Case study #2 in the main text.

| Case | Tumour type | Category | WGS genomic finding | RNAsum finding | Clinical significance |
|---|---|---|---|---|---|
| Case 1 | Adenoid cystic carcinoma | Diagnosis support | Genomic rearrangement involving NFIB and a region ~52 kb upstream of MYB | High MYB expression (99th percentile vs. TCGA PANCAN cohort) | Regulatory rearrangement placing MYB under NFIB enhancer control; consistent with adenoid cystic carcinoma [PMID: 26829750] |
| Case 16 | Not specified | Diagnosis support | EWSR1::ATF1 fusion predicted; frameshift variant identified at DNA level | Exon skipping demonstrated by WTS, restoring in-frame fusion transcript | WTS-detected exon skipping resolved the predicted frameshift, confirming expression of |

| Case | Tumour type | Category | WGS genomic finding | RNAsum finding | Clinical significance |
|---|---|---|---|---|---|
| | | | | | an in-frame EWSR1::ATF1 oncogenic fusion |
| Case 19 | Not specified | Diagnosis support | Complex inter-chromosomal translocation involving ~6.7 kb JAZF1 fragment (chr7:67,486,930-67,480,253), predicting JAZF1::SUZ12 in-frame fusion | In-frame oncogenic fusion transcript (JAZF1 exon 3 - SUZ12 exon 2) confirmed by WTS | WTS confirmed in-frame fusion arising from a complex genomic rearrangement, resolving ambiguity about the predicted fusion reading frame |
| Case 39 | Diffuse large B-cell lymphoma (DLBCL) | Diagnosis support | Limited genomic evidence of IGH::BCL6 translocation | IGH::BCL6 fusion transcript detected by WTS | WTS provided fusion evidence absent from WGS alone; IGH::BCL6 rearrangement consistent with DLBCL [PMID: 29713087, 26702065] |
| Case 43 | Hormone receptor-positive metastatic breast cancer | Treatment decision | Complex interchromosomal translocation, predicted to generate ESR1::PLEKHG1 fusion gene. Breakpoints between ESR1 ENST00000440973 exons 4 and 5, and between PLEKHG1 ENST00000358517 exons 11 and 12. Whole | In-frame fusion transcript coupling ESR1 exon 4 to PLEKHG1 exon 12 confirmed by WTS | ESR1 exon 4 fusion may predict resistance to ER degrader therapy; WTS clarified complex rearrangement to inform endocrine treatment selection |

| Case | Tumour type | Category | WGS genomic finding | RNAsum finding | Clinical significance |
|---|---|---|---|---|---|
| | | | transcriptome sequencing detected expression of an in-frame fusion transcript coupling ESR1 exon 4 to PLEKHG1 exon 12. | | |
| Case 47 | Pheochromocytoma | Treatment decision | Interchromosomal translocation (chr10:43,128,517 within RET exon 20; chr17:77,474,756 between SEPTIN9 exons 3-4) predicting RET::SEPTIN9 in-frame fusion | In-frame fusion transcript coupling RET exon 19 to SEPTIN9 exon 4 confirmed by WTS | Novel RET fusion confirmed by WTS; led to successful treatment with RET inhibitor selpercatinib [PMID: 33539223] |
| Case 49 | Breast cancer | Treatment decision | ZHX3::BRCA1 gene fusion detected | Fusion transcript involving full-length BRCA1 ORF detected; high BRCA1 expression (94th percentile vs. TCGA BRCA cohort) | Rearrangement placing BRCA1 under a heterologous promoter; known mechanism of treatment resistance in BRCA1 promoter hypermethylation-driven breast cancer [PMID: 27381626] |
| Case 55 | Breast cancer | Treatment decision | SINHCAF::ABCB1 gene fusion detected | Fusion transcripts involving full-length ABCB1 ORF detected; very high ABCB1 expression (99th | Rearrangement placing ABCB1 under a strong heterologous promoter; known mechanism of chemotherapy resistance |

| Case | Tumour type | Category | WGS genomic finding | RNAsum finding | Clinical significance |
| --- | --- | --- | --- | --- | --- |
| | | | | percentile vs. TCGA BRCA cohort) | [PMID: 30894541, 26017449] |
